# Top-Down Proteomics in the Assessment of Kidney Donor Quality: a novel approach to increased organ utilization

**DOI:** 10.64898/2026.02.02.26345404

**Authors:** Claudia Ctortecka, Dinesh Jaishankar, Pei Su, Che-Fan Huang, Indira Pla, Nathaniel Henning, Michael A. R. Hollas, Michelle A. Callegari, Meredith E. Taylor, Yu Min Lee, Amna Daud, David F. Pinelli, Vinayak Rohan, Michael A. Caldwell, Eleonora Forte, Aniel Sanchez, Neil L. Kelleher, Satish N. Nadig

## Abstract

Kidney transplantation faces a critical paradox: while thousands await organs, approximately 30% of potential deceased donor kidneys are discarded for various reasons, including *subjective* assessments due to the lack of an objective molecular biomarker of preservation quality. Here, we applied novel “top-down” proteoform imaging mass spectrometry across living donor (LD), deceased donor (brain death or cardiac death), and discarded human kidneys to quantify proteoforms correlating with post-transplant kidney function. This approach preserves post-translational modifications and splice variants, revealing molecular tissue variability beyond protein presence.

LD kidneys displayed robust metabolic signatures, including L-xylulose reductase and cytochrome oxidase subunits, whereas deceased donor and discarded organs showed elevated cellular stress markers such as alpha-B-crystallin and peroxiredoxin 1. Post-transplant blood proteoform analysis validated tissue findings, demonstrating persistent cellular stress and immune activation in deceased donor recipients compared with physiologic wound healing in LD recipients. Consistent with these molecular predictions, serum creatinine levels were highest in DCD, intermediate in DBD, and lowest in LD recipients.

The intersection of tissue proteoform signatures across all marginal tissues identified four proteoforms consistently elevated in deceased and discarded kidneys: ACTG1, acetylated CRYAB, PARK7, and S100A4. Collectively, these proteoforms capture key molecular indicators of graft quality, reflecting oxidative stress, cellular injury, and immune activation pathways. As such, they represent promising point-of-care (POC) biomarker candidates for *objective* kidney classification, potentially improving donor kidney utilization.

**Translational statement:** Current methods for evaluating donor kidney quality rely on subjective assessments, contributing to the discard of approximately 30% of potentially viable organs. This study demonstrates that “top-down” proteomics can objectively identify molecular signatures distinguishing high-quality from marginal donor kidneys. Top-down proteomics analyzes intact proteins with their post-translational modifications or cleavage products, termed proteoforms to provide mechanistic insights into graft quality. We identified four proteoforms (ACTG1, acetylated CRYAB, PARK7, and S100A4) to be consistently elevated in deceased and discarded kidneys, reflecting oxidative stress, cellular injury, and immune activation. These molecular markers correlated with post-transplant kidney outcome, as measured by serum creatinine levels and recipient blood proteoforms. As a next step, validation in larger cohorts could establish these proteoforms as point-of-care biomarkers for real-time donor kidney assessment during procurement. This objective molecular stratification could reduce unnecessary organ discards and improve transplant outcomes by matching organ quality with recipient risk profiles.

## Introduction

Kidney transplantation (KTx) is the only available curative option for patients suffering from end-stage renal disease. Yet, there is a significant disparity between the availability of donor kidneys and the number of patients requiring KTx. According to the United States Organ Procurement and Transplantation Network, as of late 2025, more than 94,000 people are on the waitlist for a KTx^1^. Despite the increased utilization of deceased donors, both from donors after brain death (DBD) and donors after cardiac death (DCD), to bridge the supply-demand gap, ∼28% of the procured kidneys are discarded^2^. Additionally, current allocation policies often result in long cold ischemic times for deceased donor kidneys. This increases the risk of slow and delayed graft function post-KTx and exacerbates ischemia-reperfusion injury, which poses a significant barrier to renal allograft longevity^3^. Thus, improving donor organ utilization rates would simultaneously bridge the supply-demand gap and enhance KTx and patient outcomes.

Since the onset of KTx in 1954, traditional metrics for donor kidneys evaluation has relied on donor and recipient medical history and demographics, histopathology of biopsies, clinical parameters such as donor urine output, serum creatinine (sCr) and estimated glomerular filtration rate (eGFR), machine perfusion parameters such as resistive indices and flow rates, and gross inspection of the renal allograft^4,5^. While these parameters affect whether kidneys are discarded or used for transplantation, no standardized clinical assessment is currently available, and many criteria are institution- and physician-dependent. This is particularly true when biopsies from DBD or DCD kidneys are deemed viable but marginally based on varying degrees of interstitial fibrosis and tubular atrophy, vascular disease, diabetic changes, or glomerulosclerosis - to name a few. Moreover, many of the traditional metrics are subjective, may not be predictive of KTx outcomes, and do not fully capture the adverse multifactorial injury processes associated with ischemia, reperfusion, and deceased donation, resulting in their detection only after transplant and irreversible injury has occurred. These limitations underscore an unmet need to develop robust transplant-relevant point-of-care (POC) molecular markers that accurately assess donor kidney quality to improve tissue utilization and mitigate post-KTx complications.

Molecular “omics” technologies have leveraged transcriptomics, proteomics, and metabolomics to objectively assess kidney quality by complementing histopathology with donor-specific molecular signatures. However, no predictive biomarker for donor kidney quality has been successfully identified. To this end, we hypothesized that top-down proteomics (TDP) may provide reliable molecular signatures undetected by ‘standard’ proteomics technologies^6,7^. TDP detects intact proteoforms, which represent molecular forms of proteins, including genetic variations in protein-coding regions, post-translational modifications (PTMs), and isoforms arising from alternative splicing or translational start sites^7,8^. Because they are more proximal to the phenotype, identification of intact proteoforms is expected to correlate more closely with the pathophysiological state of a donor organ during procurement and preservation, thereby providing an objective and reliable assessment of the donor kidney’s quality^9^. To date, no study has examined pre-transplant proteomic signatures as a measure of renal allograft quality.

In this study, we implemented TDP technology to identify proteoforms and pathways associated with donor kidney quality. Specifically, we probed the proteoform landscapes of human LD, DBD, and DCD kidney biopsies using proteoform imaging mass spectrometry (PiMS) and systematically investigated their impact on recipient peripheral blood mononuclear cells (PBMCs) post-KTx. We identified proteoform signatures associated with cellular stress and immune cell activation in kidney tissue biopsies and in PBMCs from recipients transplanted with deceased-donor kidneys. In contrast, we found increased metabolic regulation in donor biopsies and blood from LD recipients. These results align with clinical knowledge of LD, DBD, and DCD kidneys and different KTx outcomes, providing a proof-of-concept for evaluating proteoforms to assess donor kidney quality. When combined with traditional metrics, these techniques could significantly improve organ utilization and post-KTx outcomes.

## Methods

### Study design

The limited tissue availability challenges in-depth proteoform characterization of clinical biopsies. Proteoform imaging mass spectrometry (PiMS) technology enables spatially resolved proteoform detection from quantity-limited tissue biopsies through the combination of nanospray desorption electrospray ionization (nano-DESI) and individual ion mass spectrometry^10,11^. We performed PiMS on high-quality biopsies from a total of 3 biological replicates of LD and low-quality discarded kidneys. In particular, we performed nano-DESI line scans on 10 µm thick cryosections, sampling multiply charged intact proteoform ions distributed across a large range of charge states.

By assigning charges directly to individual ion signals, we generated proteoform profiles for each pixel in the imaging data. These proteoform patterns were queried for known intact proteoform masses, including phosphorylation, acetylation, and methylation.

Additionally, a total of 3 biological replicates per LD: n = 3, DBD: n = 3, or DCD: n = 3 were analyzed using PiMS to establish proteoform landscapes of high and lower quality tissues that still represent KTx candidates.

Whole blood samples of 5 biological replicates were enriched for PBMCs for a total of 15 recipients, in part overlapping with the PiMS donor samples (LD: n = 5; DBD: n = 5; DCD: n = 5). Samples were collected at two time points, immediately pre-KTx and 7 days post-KTx, then enriched for PBMCs and subjected to liquid chromatography tandem mass spectrometry (LC-MS/MS) in technical triplicates for proteoform detection. PBMC proteoforms describe the effect of the distinct kidney groups (LD, DBD, or DCD) on the recipients. For standard clinical metabolic testing, whole blood samples were obtained through year one post-KTx at the following timepoints: immediately pre-KTx, day 7, day 14, day 30, day 60, day 90, month 6, month 9, and month 12 post-KTx. Metabolic testing included sCr, eGFR, and blood urea nitrogen (BUN).

### Cohort Information

KTx recipients were enrolled in the trial at time of transplant and included adult males or females undergoing primary or subsequent deceased donor (DD) or LD KTx. Patients with a history of non-renal transplants, active immunosuppression (IS) therapy, or requiring desensitization prior to KTx were excluded. The study included LD recipients and standard grade DD kidneys, defined as those with a Kidney Profile Index (KDPI) of 30% to 80%. Demographic and clinical information for donors and recipients enrolled in this study are presented in **Table 1**. The study period spanned from 3/10/2023 to 4/20/2024.

**Table 1:** Recipient and donor cohort demographics and characteristics.

|  | Overall<br>N = 15 | DBD<br>N = 5 | DCD<br>N = 5 | Living<br>N = 5 | *p-value* | Overall<br>N = 9 | DBD<br>N = 3 | DCD<br>N = 3 | Living<br>N = 3 | *p-value* |
| --- | --- | --- | --- | --- | --- | --- | --- | --- | --- | --- |
| **Recipient Characteristics** |  |  |  |  |  |  |  |  |  |  |
|  | PBMC |  |  |  |  | PiMS |  |  |  |  |
| Age, years | 60 (52, 61) | 58 (52, 60) | 59 (53, 60) | 61 (60, 61) | 0.6 | 53 (50, 60) | 50 (45, 55) | 53 (52, 59) | 60 (53, 61) | 0.575 |
| Male | 11 (73%) | 3 (60%) | 4 (80%) | 4 (80%) | >0.9 | 8 (89%) | 3 (100%) | 3 (100%) | 2 (67%) | >0.9 |
| Race |  |  |  |  | 0.3 |  |  |  |  | 0.4 |
| Black or African American | 5 (33%) | 2 (40%) | 3 (60%) | 0 |  | 4 (44%) | 2 (67%) | 2 (67%) | 0 |  |
| White | 10 (67%) | 3 (60%) | 2 (40%) | 5 (100%) |  | 5 (56%) | 1 (33%) | 1 (33%) | 3 (100%) |  |
| Ethnicity |  |  |  |  | >0.9 |  |  |  |  |  |
| Hispanic/Latino | 1 (6.7%) | 1 (20%) | 0 |  |  |  |  |  |  |  |
| Not Hispanic/Latino | 14 (93%) | 4 (80%) | 5 (100%) | 5 (100%) |  | 9 (100%) | 3 (100%) | 3 (100%) | 3 (100%) |  |
| Blood Type |  |  |  |  | 0.14 |  |  |  |  | >0.9 |
| A | 7 (47%) | 2 (40%) | 1 (20%) | 4 (80%) |  | 4 (44%) | 1 (33%) | 1 (33%) | 2 (67%) |  |
| AB | 1 (6.7%) | 0 | 1 (20%) | 0 |  | 2 (22%) | 1 (33%) | 1 (33%) | 0 |  |
| B | 1 (6.7%) | 0 | 0 | 1 (20%) |  | 1 (11%) | 0 | 0 | 1 (33%) |  |
| O | 6 (40%) | 3 (60%) | 3 (60%) | 0 |  | 2 (22%) | 1 (33%) | 1 (33%) | 0 |  |
| Etiology of Kidney Disease |  |  |  |  | 0.7 |  |  |  |  | 0.2 |
| Diabetes Mellitus | 5 (33%) | 1 (20%) | 3 (60%) | 1 (20%) |  | 5 (56%) | 1 (33%) | 3 (100%) | 1 (33%) |  |
| Glomerulonephritis | 2 (13%) | 1 (20%) | 1 (20%) | 0 |  |  |  |  |  |  |
| Hypertensive Nephrosclerosis | 1 (6.7%) | 1 (20%) | 0 | 0 |  | 1 (11%) | 1 (33%) | 0 | 0 |  |
| Polycystic Kidney Disease | 3 (20%) | 1 (20%) | 0 | 2 (40%) |  | 2 (22%) | 0 | 0 | 2 (67%) |  |
| Other/Unknown | 4 (27%) | 1 (20%) | 1 (20%) | 2 (40%) |  | 1 (11%) | 1 (33%) | 0 | 0 |  |
| History of Kidney Transplant | 2 (13%) | 2 (40%) | 0 | 0 | 0.3 | 1 (11%) | 1 (33%) | 0 | 0 | >0.9 |
| Pre-Transplant Dialysis Type |  |  |  |  | 0.2 |  |  |  |  | >0.9 |
| Hemodialysis | 9 (60%) | 4 (80%) | 4 (80%) | 1 (20%) |  | 5 (56%) | 2 (67%) | 2 (67%) | 1 (33%) |  |
| Peritoneal Dialysis | 2 (13%) | 1 (20%) | 0 | 1 (20%) |  | 2 (22%) | 1 (33%) | 0 | 1 (33%) |  |
| None | 4 (27%) | 0 | 1 (20%) | 3 (60%) |  | 2 (22%) | 0 | 1 (33%) | 1 (33%) |  |
| Total Waiting Time, days<sup>a</sup> | 1,075 (291, 1,913) | 1,913 (1,753, 1,921) | 1,075 (881, 1,286) | 291 (209, 319) | 0.01 |  |  |  |  |  |
| Time on Dialysis, years | 3.0 (1.0, 3.7) | 1.0 (1.0, 4.9) | 3.3 (2.9, 3.6) | 1.6 (0.8, 2.3) | 0.5 | 2.7 (1.8, 3.4) | 2.7 (1.9, 3.8) | 3.2 (2.9, 3.5) | 1.6 (0.8, 2.3) | 0.738 |
| **Donor Characteristics** |  |  |  |  |  |  |  |  |  |  |
|  | PBMC |  |  |  |  | PiMS |  |  |  |  |
| Age, years | 51 (37, 58) | 50 (50, 53) | 60 (53, 60) | 38 (35, 51) | 0.13 | 51 (45, 58) | 50 (48, 54) | 60 (57, 60) | 35 (34, 43) | 0.078 |
| Male | 8 (53%) | 2 (40%) | 4 (80%) | 2 (40%) | 0.5 | 6 (67%) | 2 (67%) | 3 (100%) | 1 (33%) | 0.7 |
| Race |  |  |  |  | 0.066 |  |  |  |  | >0.9 |
| Asian | 1 (6.7%) | 1 (20%) | 0 | 0 |  | 1 (11%) | 1 (33%) | 0 | 0 |  |
| Black or African American | 1 (6.7%) | 1 (20%) | 0 | 0 |  |  |  |  |  |  |
| Other | 1 (6.7%) | 1 (20%) | 0 | 0 |  |  |  |  |  |  |
| White | 12 (80%) | 2 (40%) | 5 (100%) | 5 (100%) |  | 8 (89%) | 2 (67%) | 3 (100%) | 3 (100%) |  |
| Ethnicity |  |  |  |  | >0.9 |  |  |  |  | >0.9 |
| Hispanic/Latino | 2 (13%) | 1 (20%) | 0 | 1 (20%) |  | 1 (11%) | 0 | 0 | 1 (33%) |  |
| Not Hispanic/Latino | 13 (87%) | 4 (80%) | 5 (100%) | 4 (80%) |  | 8 (89%) | 3 (100%) | 3 (100%) | 2 (67%) |  |
| Blood Type |  |  |  |  | >0.9 |  |  |  |  | >0.9 |
| A | 4 (27%) | 2 (40%) | 1 (20%) | 1 (20%) |  | 3 (33%) | 1 (33%) | 1 (33%) | 1 (33%) |  |
| AB | 1 (6.7%) | 0 | 1 (20%) | 0 |  | 2 (22%) | 1 (33%) | 1 (33%) | 0 |  |
| O | 10 (67%) | 3 (60%) | 3 (60%) | 4 (80%) |  | 4 (44%) | 1 (33%) | 1 (33%) | 2 (67%) |  |

The research protocol was approved by Northwestern Medicine Institutional Review Board (IRB). All the patients in this study consented to the use of their anonymized data, and their data are summarized in **Table 1**.

### Patient sample collection and processing

#### Tissue Biopsies

Back-table kidney tissue biopsies were collected intraoperatively by the surgical team at Northwestern Medicine Comprehensive Transplant Center. Wedge tissue biopsies were acquired from LD, DBD, or DCD donors. All biopsies were obtained from isolated kidney allografts before implantation and were processed as follows: each biopsy was obtained from the upper pole of the kidney cortex, snap-frozen in liquid nitrogen-cooled isopentane, and stored in a cryotube at −196°C before further processing and analysis. Additionally, three wedge biopsies from discarded kidneys (low quality and unsuitable for transplant) were included in this study.

Cryostat (Leica) was conditioned to −16°C to obtain cryosections (10 µm), from which five high-quality adjacent cryosections were selected for each biopsy. Each section was thaw-mounted onto a positively charged glass microscope slide (Globe Scientific) and stored at −80 °C before PiMS analysis. Optical images of the tissue sections used for PiMS analysis are shown in **Supplementary Figures S1, S2 and S3.**

For PiMS sample pretreatment, the tissue sections were thawed at room temperature under slight vacuum in a desiccator, fixed and desalted via successive immersion in 70%/30%, 90%/10%, and 100%/0% (*v*/*v*) ethanol/water solutions for 20 s each, followed by a delipidation step using 99.8% chloroform for 60 s, and dried at room temperature. As a final step prior to the PiMS experiment, the tissue sections were optically scanned using a PathScan Enabler (Thermo Fisher Scientific, Waltham, MA).

### Blood Samples

Blood samples from recipient patients were collected at two-time points. Pre-KTx (day 0) samples were drawn prior to the administration of any IS/induction therapies. The second sample was collected on day 7 post-KTx. CPT tubes were processed within one hour of collection and spun at room temperature at 1,700 *g* for 20 minutes. The PBMC layer was collected and washed with phosphate-buffered saline, and cells were counted, diluted to a final concentration of 5 × 10^6^ cells/mL, and preserved at −196°C in 80% Fetal Bovine Serum/20% dimethyl sulfoxide. Cells were lysed in 20 mM Tris-HCl pH 7.5, 100 mM NaCl, 1% N-lauroylsarcoside, and protease/phosphatase inhibitor cocktails. Lysates were sonicated, proteins were precipitated, resuspended in 1% Sodium dodecyl sulfate, and their concentrations were determined by BCA assay. Proteins were fractionated by Passively Eluting Proteins from Polyacrylamide gels as Intact species, and proteins with a molecular weight lower than 30 kDa, as described previously^12^, were analyzed by liquid chromatography-tandem mass spectrometry (LC-MS/MS) on an Orbitrap Ascend Tribrid mass spectrometer (Thermo Fisher Scientific, San Jose, CA).

### Top-Down Liquid Chromatography-Mass Spectrometry

Protein samples were separated using a Vanquish Neo UHPLC system (Thermo Fisher Scientific) as previously described^13,14^. Briefly, reversed-phase liquid chromatography was conducted on a MAbPac EASY-Spray column (150 mm length by 150 µm i.d., Thermo Fisher Scientific) and an in-house packed PLRP-S trap column (25 mm length by 150 µm i.d., Agilent). The total run time was 120 minutes, utilizing a gradient of mobile phase A (99.9% water, 0.1% FA) and mobile phase B (19.9% water, 80% acetonitrile 0.1% Formic Acid). The flow rate was maintained at 1 µL/min. The gradient profile was as follows: 5% B at 0 minutes, 20% B at 5 minutes, 70% B at 110 minutes, 99% B from 111 to 114 minutes, and 5% B from 115 to 120 minutes. The column outlet was connected in line to an EASY-Spray source on an Orbitrap Ascend mass spectrometer (Thermo Fisher Scientific) operating in intact protein mode with 2 mTorr of N_2_ pressure in the ion routing multipole. The transfer capillary temperature was set to 320°C, the ion funnel RF was set to 60%, and a source collision induced dissociation (CID) of 15 Volts was applied. MS1 spectra were acquired at resolution of 120,000 power (at m/z 200), normalized AGC target of 500%, maximum injection time 100ms and 1 µscan. Target precursors were isolated using a 3 m/z window for data-dependent topN MS2 method included 60,000 (at m/z 200), resolution, 27 NCE for HCD with a normalized AGC of 800%, 800 ms maximum injection time and 1 µscan with a dynamic exclusion of 60s and a threshold of 1x104 intensity

### PiMS ion source and sampling conditions

PiMS utilizes nanospray desorption electrospray ionization MS coupled with Orbitrap-based individual ion MS (I^2^MS), which has been described in detail elsewhere^15,16^. The nano-DESI probe is comprised of two capillaries: a primary capillary that delivers extraction solvent to the tissue, a nanospray capillary for transfer and ionization of extracted proteoforms, interfacing with mass spectrometry (MS) detection^16^. Localized analyte extraction from tissue is sampled by a dynamic liquid bridge formed between the junction of the two capillaries and the tissue section. In these experiments, the tissue sample was moved under the PiMS probe at a raster scanning rate of 4 µm/s with a strip step of 100 µm, which allows proteoforms from different spatial locations to be sampled and transferred to MS for detection as individual ions^11^. In all PiMS experiments, a denaturing extraction solvent (60%/39.4%/0.6% (*v*/*v/v*) acetonitrile/water/glacial acetic acid) was used at a flow rate of 400 nL/minute, allowing efficient proteoform extraction and ionization in positive mode.

The PiMS probe used in this study was fabricated using fused silica capillaries (Molex, Thief River Falls, MN, OD/ID 150/40 µm). To improve sampling sensitivity and spatial resolution, the primary capillary was flame-pulled to an OD of 20 µm on one side to reduce the liquid bridge diameter. A three-point plane was defined in the tissue section to compensate for the tilting angle, ensuring the stability of the liquid bridge during spatial profiling. For each tissue biopsy section, a continuous tissue area of approximately 4 mm^2^ was selected as the region of interest for PiMS spatial proteoform profiling. The optical images of the exact areas selected for each biopsy are shown in **Supplementary Figure S1** (LD), **Supplementary Figure S2** (DBD), and **Supplementary Figure S3** (DCD).

### PiMS spatial data acquisition and processing

PiMS experiments were conducted on the Orbitrap Exploris 480 mass spectrometer (Thermo Fisher Scientific, Bremen, Germany) in the I^2^MS mode at a resolution of 120,000 at *m/z* 200, which corresponds to 0.5 s Orbitrap detection period (a data acquisition rate of 2 spectra/s)^17^. The source conditions for the mass spectrometer were set as follows: electrospray ionization voltage: 3 kV; in-source CID: 5 eV; S-Lens/Funnel RF level: 70%; capillary temperature: 325°C. MS injection time was kept at 0.2 milliseconds to minimize “multi-ion” signals with overlapping *m/z* values from multiple individual ions in the same detection period. The injection time has been systematically optimized and benchmarked using a spare kidney tissue section from the same sample set. Other instrument parameters used in these experiments were listed as follows: Higher-energy collisional dissociation pressure setting: 0.33 (arbitrary unit); extended trapping: 0.5 V; mass range: 400-2500 *m/z*; AGC mode: fixed; enhanced Fourier transform: off; Emeter averaging: 0; microscans: 1.

PiMS data acquisition was performed in the I^2^MS mode enabled by a Developers license as described in detail elsewhere^10^. In short, individual ions from the tissue were collected by the PiMS probe as a function of location; time-domain data files were acquired and recorded as Selective Temporal Overview of Resonant Ions (STORI) files^18^. Proteoform features were extracted by a modified THRASH algorithm tailored for the I^2^MS datatype represented by their monoisotopic masses and ion counts^19^.

### PiMS proteoform identification and quantitation

Proteoform identification was performed using an Intact Mass Tag (IMT) approach described in detail previously^10^. Specifically, I^2^MS mass spectrum of each biopsy was first systematically calibrated according to the accurate masses of six known abundant kidney proteoforms in the 4-50 kDa mass range. The calibrated spectrum was subsequently converted to .mzML format, and the proteoform features in the spectrum were searched against a custom proteoform database using TDValidator (Proteinaceous, Evanston, IL) implemented with an MS^1^ IMT search function with a ±1.5 ppm mass tolerance for matching. In particular, the proteoform database comprises 259 highly curated kidney proteoforms from a previous PiMS dataset, acquired from healthy human kidney tissues. Abundances of identified proteoforms were extracted as ion counts and normalized to the total ion count (TIC) in each spectrum for subsequent analysis.

Using RStudio platform^20^, TIC normalized proteoform abundances were subjected to a nested ANOVA analysis to measure and significance in differential proteoform expression among the three patient groups. Unsupervised hierarchical clustering of normalized proteoform abundances was performed using ‘ComplexHeatmap’ (version 2.18.0)^21^ R package. For both, fold-changes of normalized proteoform data LD versus DBD/DCD were calculated and subjected to pathway analysis within Reactome (https://reactome.org/). Vulcano, boxplots, bar graphs, density plots and false discovery rate (FDR) filtered pathways per grouped comparison were plotted using ‘ggplot2’ (version 4.0.0) R package with highlights of immune defense, cell stress/cell death, and glucose metabolism, or previously identified proteoforms^8^.

### LC-MS/MS data processing and analysis

The raw data files were analyzed using the publicly accessible standard workflow on TDPortal (https://portal.nrtdp.northwestern.edu, Code Set 4.0.0). This workflow conducts mass inference and searches against a database of human proteoforms, derived from SwissProt (June 2020), with curated histones, maintaining a 1% FDR at the protein, isoform, and proteoform levels^22^. A proteoform database was created from SwissProt containing 2.4 million proteoform entries. For quantitative proteoform analysis, a CSV intensity sheet was generated using an isotopic fitting algorithm across the chromatogram to determine the intensities of all proteoforms identified in the study with 10% FDR confidence in each sample. This included those previously classified as ‘unidentified’ and not meeting the 1% FDR identification threshold^23^. The three initial searches used the same parameters and search space. Therefore, the intermediate search result files were combined (.pufdb) into a single .tdReport, and a 1% FDR threshold was applied across all files.

Statistical analysis were performed using the RStudio platform^20^. Data derived from randomized biological and technical replicates were used to assess analytical performance and differential proteoform expression among the patient groups. Missing values were filtered to keep proteoforms quantified in at least 50 percent of the samples from at least one biological condition (i.e., group). To stabilize variance and make the data more comparable across samples, intensities were Log_2_ transformed and normalized by subtracting the median intensity of each sample from all its intensities and then dividing the result by the sample standard deviation. To assess differentially expressed proteoforms, the log_2_-transformed intensities were z-score normalized across samples and used as a dependent variable in a hierarchical mixed linear regression model created using the ‘lme4’ (version 1.1.38) R package^24^. Biological and technical replicates were set as random nested variants in the model, and p-values were adjusted using the Benjamini and Hochberg approach to control the FDR induced by multiple testing^25^. In all cases, adjusted p-values <0.05 were considered significant. Volcano plots were generated to display, for each proteoform, the statistical confidence of the difference between two groups (-log_10_ of adjusted p-value) as a function of its fold change (log_2_ fold-change). Gene ontology (GO) analyses were performed on the Metascape online tool (https://metascape.org//)^15^. For ease of display the following Reactome pathways were group to ‘*Cell stress & death*’: Cellular response to chemical stress, Cellular responses to stimuli, Cellular responses to stress, Chaperone Mediated Autophagy, Late endosomal microautophagy, Selective autophagy, Defective Intrinsic Pathway for Apoptosis, Apoptosis, Regulation of Apoptosis, Apoptosis induced DNA fragmentation, Macroautophagy, Autophagy, Regulation of MITF-M-dependent genes involved in lysosome biogenesis and autophagy, FOXO-mediated transcription of oxidative stress, metabolic and neuronal genes, Cellular response to heat stress, Regulation of MITF-M-dependent genes involved in apoptosis, Turbulent (oscillatory, disturbed) flow shear stress activates signaling by PIEZO1 and integrins in endothelial cells, Intrinsic Pathway for Apoptosis, High laminar flow shear stress activates signaling by PIEZO1 and PECAM1:CDH5:KDR in endothelial cells, Response of endothelial cells to shear stress, Cellular responses to mechanical stimuli; *Metabolism*: Aerobic respiration and respiratory electron transport, Metabolism, TP53 Regulates Metabolic Genes, Glucose metabolism, Manipulation of host energy metabolism, Metabolism of carbohydrates and carbohydrate derivatives, Glycogen metabolism, Pyruvate metabolism, Regulation of pyruvate metabolism, Diseases of carbohydrate metabolism, Selenoamino acid metabolism, Metabolism of proteins, Metabolism of porphyrins, Biotin transport and metabolism, Phenylalanine metabolism, Metabolism of amino acids and derivatives, Vitamin B5 (pantothenate) metabolism, Phenylalanine and tyrosine metabolism, Metabolism of nitric oxide: NOS3 activation and regulation, Diseases of metabolism, Glyoxylate metabolism and glycine degradation, Metabolism of RNA, Triglyceride metabolism, Regulation of lipid metabolism by PPARalpha, Fatty acid metabolism, Metabolic disorders of biological oxidation enzymes, Sulfur amino acid metabolism, PI Metabolism, Metabolism of nucleotides, Arachidonate metabolism, Metabolism of water-soluble vitamins and cofactors, Phospholipid metabolism, Metabolism of vitamins and cofactors, Metabolism of lipids; *Immune Defense*: Gene and protein expression by JAK-STAT signaling after Interleukin-12 stimulation, Antigen processing-Cross presentation, Innate Immune System, MHC class II antigen presentation, Antigen processing: Ubiquitination & Proteasome degradation, Class I MHC mediated antigen processing & presentation, Adaptive Immune System; *Platelet activation*: Platelet degranulation, Platelet activation, signaling and aggregation. Venn diagrams were performed using the BioVenn application (https://www.biovenn.nl/)^26^.

### Statistical Analysis for Clinical Data

Data on recipient and donor characteristics, transplant information, and post-transplant outcomes were summarized as medians (interquartile ranges, IQR) and counts (proportions) for continuous and categorical variables, respectively. Comparisons between DCD, DBD, and LD groups were analyzed with the Kruskal-Wallis rank sum test for continuous variables or Fisher’s exact test for categorical variables. A p-value of <0.05 was considered statistically significant. All analyses were performed in R 2024.04.2 using RStudio^20^.

## Results

### PiMS reveals molecular profiles of distinct donor kidney qualities

To evaluate whether PiMS is suitable for probing molecular definitions of donor kidney quality, we performed PiMS on biopsies from high-quality LD and low-quality discarded kidneys (**Fig. 1a**). The proteoform landscapes for three LD and three discarded kidney biopsies yielded a total of 784 proteoforms. Of these, 662 proteoforms were common to both sample types, indicating high similarity between LD and discarded groups, while 37 and 85 proteoforms were noted unique to LD and discarded biopsies, respectively (**Fig. 1b**). Interestingly, unique proteoforms in the LD are involved in NADH dehydrogenase (i.e., NDUFC2, NDUFS3, NDUFB10) or glycolysis pathways (i.e., PGAM1, ALDOA, G6PD), while unique discarded proteoforms suggest elevated activity in fatty acid metabolism (i.e., PPT1, SCP2, CBR4).

**Figure 1.**
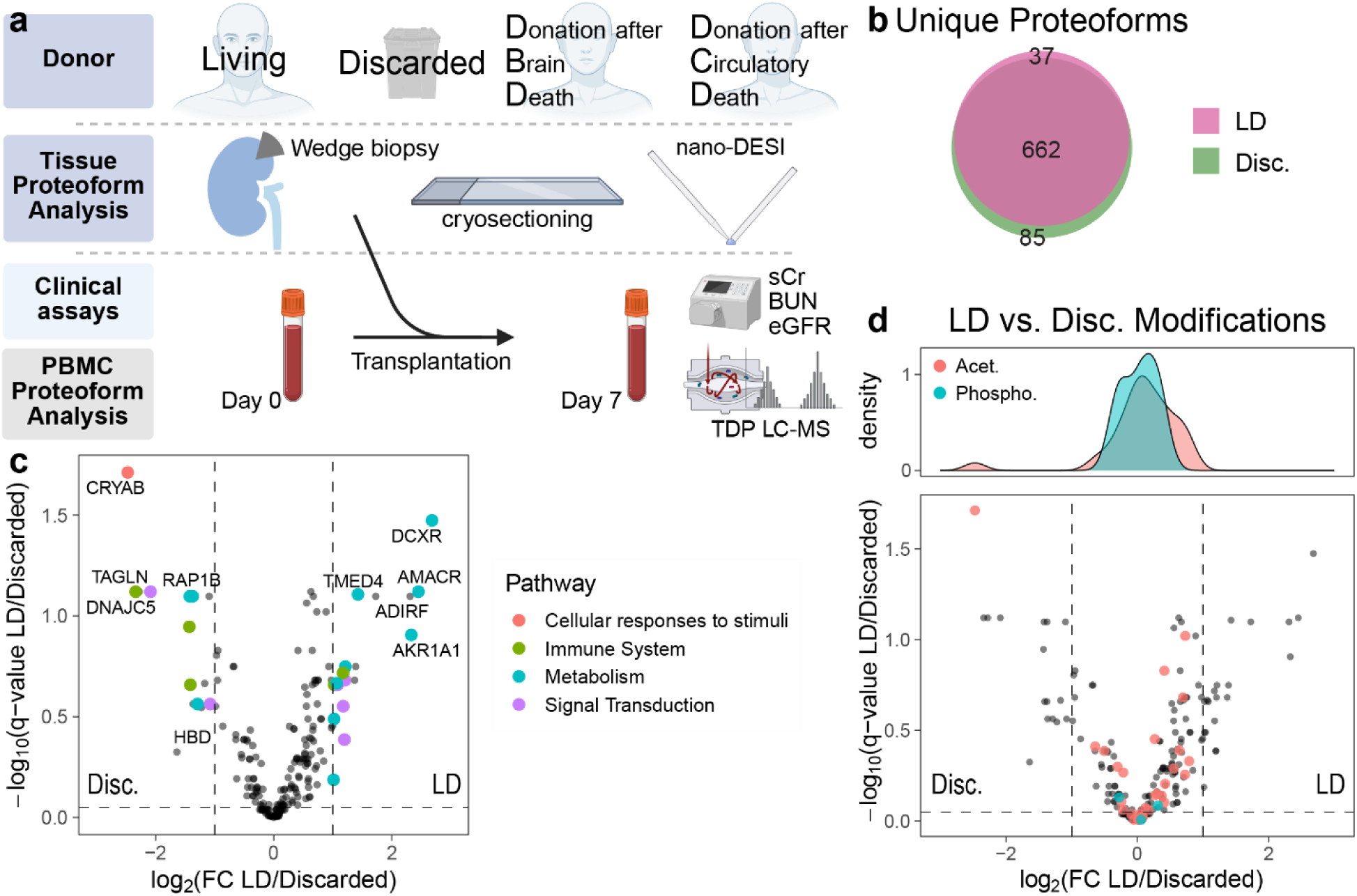
Tissue proteoform imaging reveals elevated levels of cellular stress and immune regulation in discarded versus transplanted kidneys. (a) Multi-level kidney proteoform profiling across living (LD), discarded (Disc.), Donation after Brain Death (DBD), and Donation after Circulatory Death (DCD) donor kidneys. nano-DESI = nanospray desorption electrospray ionization for direct tissue proteoform analysis; TDP LC-MS = top-down proteomics via liquid chromatography mass spectrometry for proteoform analysis. sCr = serum creatinine, BUN = blood urea nitrogen, eGFR = estimated glomerular filtration rate. Created in BioRender. https://BioRender.com/60jyi32 (b) Venn diagram of LD and discarded kidney tissue proteoforms based on PiMS analysis. (c) Log_2_ fold-change and –log_10_ q-value volcano plot of LD versus discarded proteoforms; top 4 regulated pathways are color coded and labeled with their parent proteins, and (d) acetylated (acet.) or phosphorylated (Phospho.) proteoforms are highlighted in the inset atop the volcano plot (contains the same data as in panel c).

Further inspection revealed only few proteoforms significantly differentially expressed between discarded and LD donor kidneys. Twenty proteoforms upregulated in LD kidneys were primarily metabolism-related, while 16 proteoforms significantly elevated in discarded kidneys included alpha crystallin B-chain (CRYAB), a stress response chaperone, as the most prominent (**Fig. 1c-d**). Multiple immune and signal transduction proteoforms were identified and a global PTM analysis revealed equal distribution between groups with slightly elevated acetylation in LD donor kidneys (**Fig. 1d**). Collectively, these results indicate that PiMS-enabled proteoform profiling identifies biologically relevant pathways and highlights possible molecular markers of tissue quality.

### Tissue proteoform analysis reveals differentially expressed proteoforms associated with oxidative stress in deceased donors

Next, interrogated tissue biopsies similar from transplant-suitable kidneys. For this, we included gold-standard LD donors and two deceased donors: DBD and DCD. Tissue analysis yielded 393 distinct proteoforms, of which hierarchical clustering separated LD and DCD from DBD tissues (**Fig. 2a-b**). Interestingly, patient clustering indicates progression from healthy to pathological states. The DBD donor with healthy transplant outcome clustered closest to LD and DCD donors, while one DBD and one DCD donor showed abnormal pathology clustered intermediately; the most divergent DBD kidney was rejected. LD kidneys had the most unique proteoforms (31) versus DCD (8) and DBD (4), with 157 proteoforms common to all groups. We identified 22 proteoforms common to LD and DCD, 11 common to LD and DBD, and only 4 common to DBD and DCD (**Fig. 2c**). This indicates LD and DCD donors share more unique proteoforms and similar quantitative landscapes compared to DBD donors.

**Figure 2.**
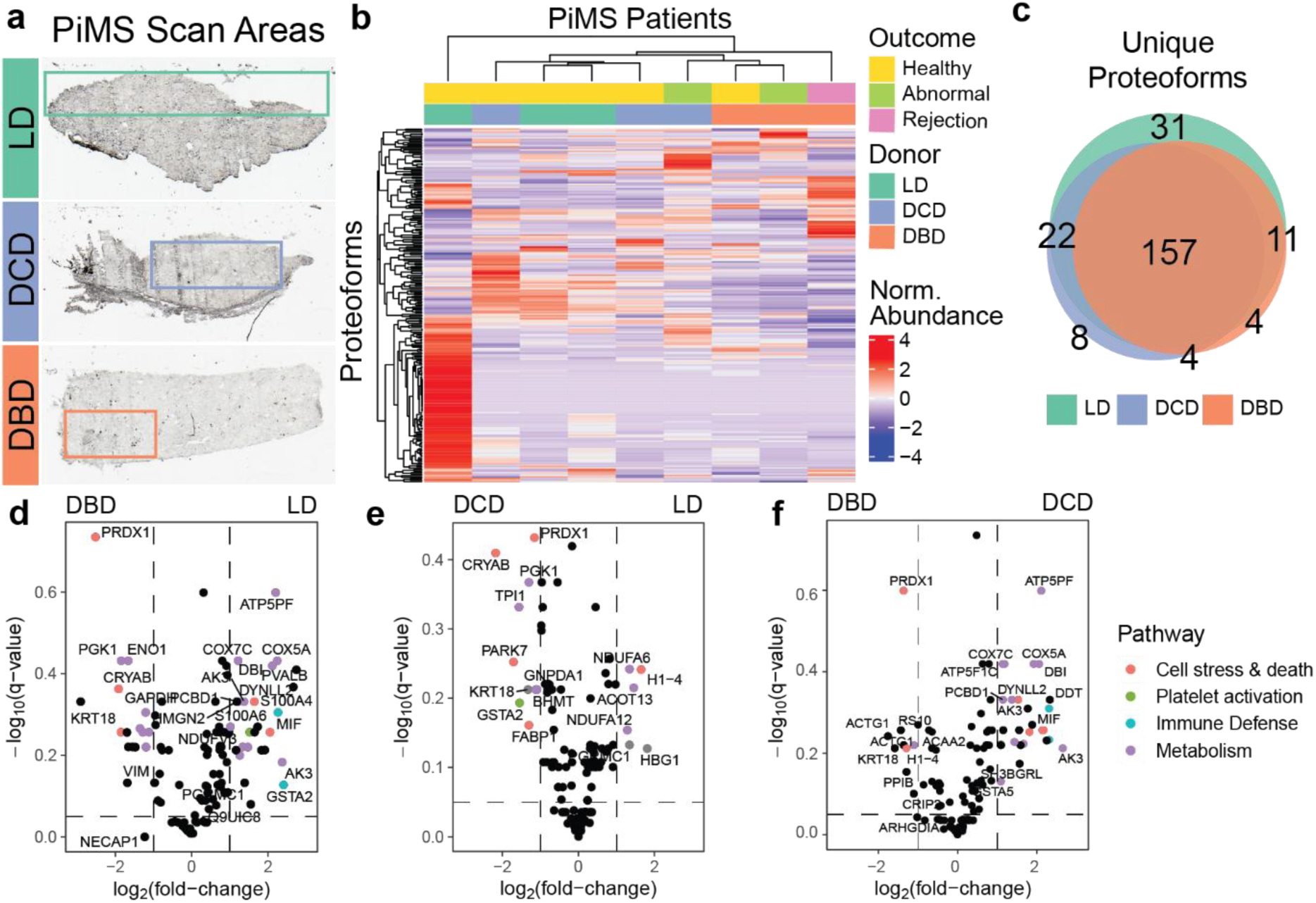
Tissue proteoforms from DCD and DBD donors indicate higher molecular similarity between DCD and LD kidneys. **(a)** Representative tissue sections with highlights of the regions acquired with PiMS. **(b)** Hierarchical clustering of normalized proteoform abundances, colored by donor group and recipient outcome. **(c)** Unique proteoform overlap for LD, DCD, and DBD donors. Log_2_ fold-change and –log_10_ q-value volcano plots of **(d)** LD versus DBD, **(e)** LD versus DCD, and **(f)** DCD versus DBD. Colors indicate representative pathways, including cell stress, platelet activation, immune defense, and metabolism (Reactome pathways detailed in Supplementary Methods). Proteoforms are labeled with their source protein.

Differential expression analysis revealed 12 and 19 proteoforms significantly more abundant in DCD and DBD, respectively, versus LD (**Fig. 2d-e**). Consistent with discarded kidneys (**Fig. 1c**), the chaperone protein CRYAB and Peroxiredoxin-1 (PRDX1), both involved in cellular protection, were elevated in deceased donor samples (**Fig. 2d-e**). Meanwhile, metabolism-related proteoforms, such as NDUFA6 and COX5A, were more abundant in the LD kidneys (**Fig. 2d-e**). Interestingly, 27 proteoforms were significantly elevated in DCD compared to DBD donors, primarily involved in cellular stress response and metabolism (**Fig. 2f**). Global comparison correlating DBD versus LD and DCD versus LD fold-changes (**Supplementary Figure 6a**) confirmed that LD grafts are metabolically superior with lower oxidative stress showing higher metabolism-involved proteoforms in LD and DCD samples.

### Recipient and Donor Characteristics for PiMS and PBMC Cohorts

Nine recipients (PiMS cohort) and 15 recipients (PBMC cohort) were enrolled at KTx (**Table 1**). Median age was 53 years (IQR 50, 60) and 60 years (IQR 52, 61), most were male (89% and 73%), respectively. The most common disease etiology was diabetes mellitus (33%, 56%, respectively). Two patients in the PBMC cohort (13% - DBD recipients) and one in the PiMS cohort (33% - DBD recipient) had previous transplants. Most patients were on dialysis pre-KTx (77.8%, 73.3%, respectively), predominantly hemodialysis versus peritoneal dialysis, with median dialysis duration of 2.7 years (IQR 1.8, 3.4) and 3 years (IQR 1.0, 3.7), respectively. Median donor age was 51 years (IQR 45-58, IQR 37-58), approximately half male. Characteristics were similar across donor types (DBD, DCD, LD) in both cohorts.

Donor kidney quality assessment (**Fig. 3a, Supplementary Figure S4)** showed median KDPI of 58% (IQR 40-63, IQR 46-70) for deceased donors. About half of deceased donor kidneys were perfused pre-implantation (56%, 47%), including all DCD kidneys in both cohorts (n=3, n=5). Resistance, flow rates (mL/minute) and cold or warm ischemic times were similar across all groups. Induction therapy was Campath or Simulect in the PiMS (78% versus 22%, respectively) and PBMC (53% versus 47%, respectively) cohorts, similar across donor types.

**Figure 3.**
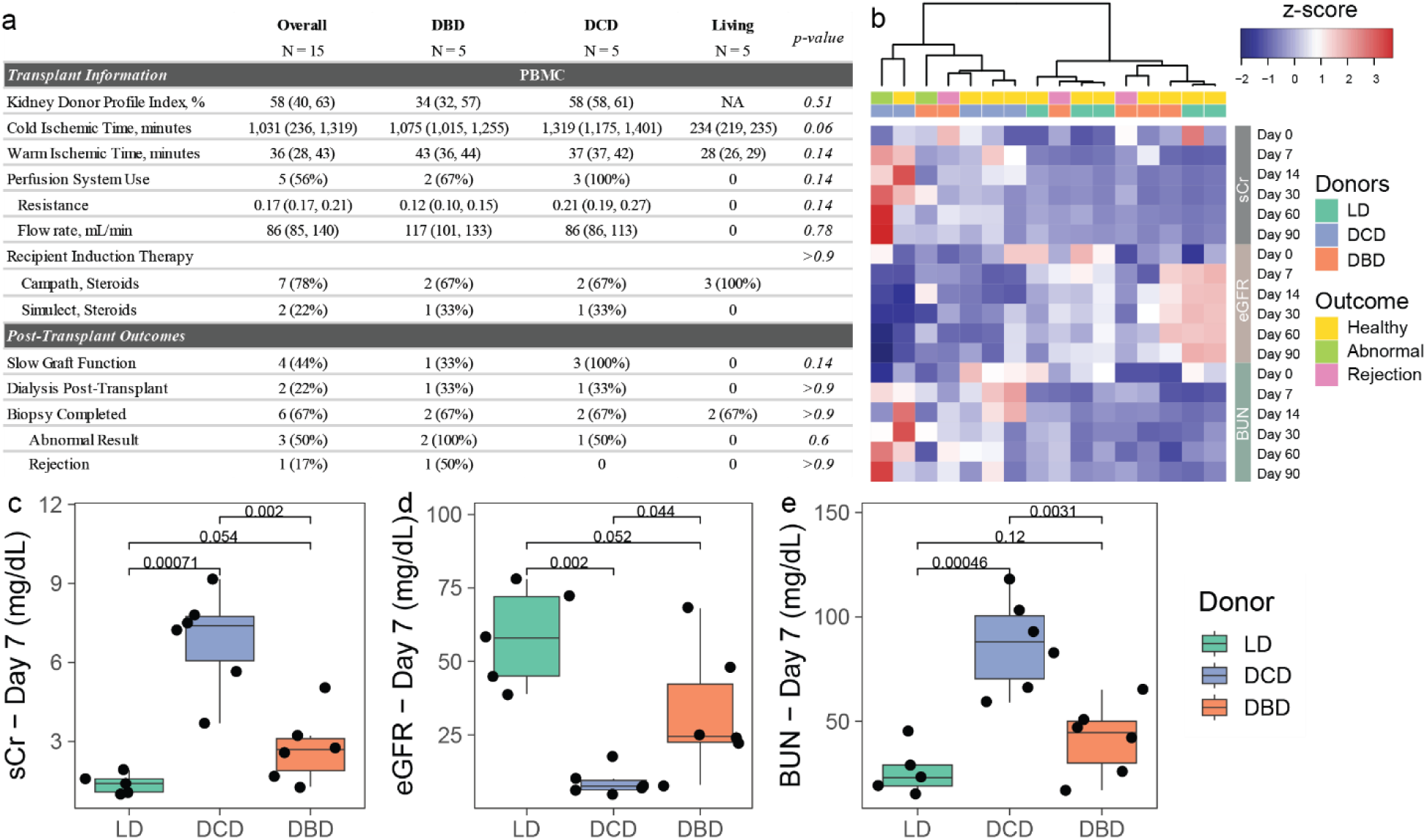
Standard clinical assays indicate higher similarities between LD and DBD recipients. **(a)** Overview of transplant and post-transplant outcomes for all recipients. **(b)** Hierarchical clustering of BUN, eGFR, and sCR levels for LD, DCD, or DBD recipients’ day 0, 7, 14, 30, 60, or 90 post-KTx. Relative comparison of **(c)** sCr, **(d)** eGFR, and **(e)** BUN levels at day 7 post-KTx for LD, DCD, or DBD recipients with ANOVA-based p-value (n = 6).

### Clinical assessment indicates a prolonged return to baseline in recipients of deceased-donor kidneys

Kidney function was monitored from pre-KTx (day 0) through 12 months post-KTx, using standard laboratory tests and clinical assessments. Standard line of care metabolic activity (i.e., sCR, BUN, and eGFR) were evaluated throughout transplant courses to assess recipient kidney function. BUN and eGFR returned more slowly to baseline in deceased donor recipients than LD donors. Hierarchical clustering of clinical values separated DCD from DBD and LD donors (**Fig. 3b**). DCD recipients showed significantly elevated creatinine levels, LD recipients indicated no elevation, and DBD recipients showed slightly higher levels 7 days post-KTx (**Fig. 3c-e**).

In the PBMC cohort, 47% were diagnosed with slow graft function (SGF)^27^, and 33% required post-KTx dialysis, both only in deceased donor recipients. The PiMS cohort showed similar early graft function outcomes. Post-KTx graft function was monitored with surveillance biopsies per institutional protocol (3 and/or 12 months) or as clinically indicated (‘For-Cause’). About one-third (PBMC 27%, PiMS 33%) did not require biopsies within the first year. Of those biopsied, half showed abnormal pathology, including moderate arteriosclerosis, primary disease recurrence, and/or tubule-interstitial cellular infiltrates (**Supplementary Table 1**). Normal results were defined as reports that did not result in immunosuppression (IS) changes. Antibody-mediated or cellular rejection was observed in 27% and 17% (n=3, n=1), respectively. DBD recipients had higher rates of abnormal pathology findings.

### Proteoforms of recipients’ blood-derived immune cells from deceased donor transplants are associated with immunological response pathways

To correlate clinical assessments with the respective proteoform landscapes, we subjected PBMC-enriched samples pre-KTx (day 0) and at day 7 post-KTx to TDP LC-MS/MS analysis. The PBMC proteoform landscape at baseline (day 0) did not cluster recipients who received LD, DBD, and DCD kidneys, as expected. However, in the day 7 samples we observed distinct separation of day 0 proteoform profiles in patients that would go on to receive DBD donor tissue and ultimately reject or show abnormal clinical outcome (**Fig. 4a**). Additionally, the fold change of blood proteoforms between day 0 and day 7 post-KTx indicated clear differences between the LD and deceased kidney recipients (**Fig. 4b**). This is supported by the large overlap of uniquely identified proteoforms between the deceased kidney recipients (2,237 or 30%), with only 253 (3.4%) or 271 (3.7%) shared between LD and DBD or DCD, respectively. Overall, we observed 3,570 total proteoforms in LD, 5,926 in DCD, and 5,702 in DBD, with 34.9% of all proteoforms across all three samples combined, but only 6.7%, 12%, or 8.7% unique to LD, DBD, or DCD donors, respectively (**Fig. 4c**).

**Figure 4.**
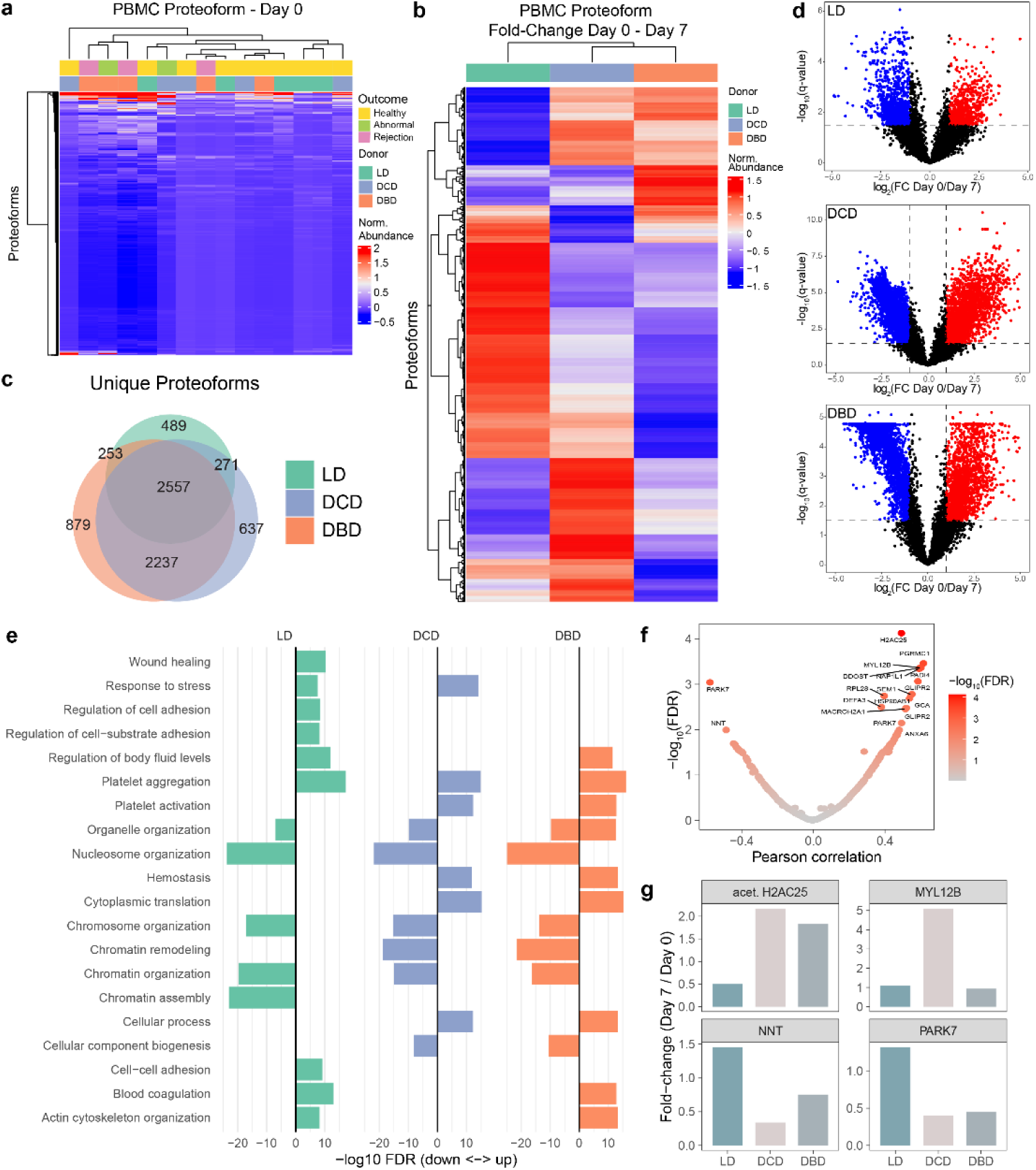
The PBMC proteoform landscape changes drastically at 7 days post-KTx. **(a)** Hierarchical clustering of PBMC proteoforms prior to KTx via LCMS analysis and **(b)** the fold change of day 0 versus day 7 for LD, DCD, or DBD recipients with the respective patients’ clinical outcome in a. **(c)** Unique proteoform overlap of LD, DCD, or DBD and **(d)** differential proteoform expression pre- and 7 days post-KTx**. (e)** Pathway enrichment analysis of significantly regulated proteoforms per recipient group (LD, DCD, and DBD). **(f)** Correlation of the blood sCr level and PBMC proteoforms on day 7. Source proteins are labeled. **(g)** Proteoform fold-changes pre- and 7 days post-KTx of the top 2 most significantly correlating (acet. H2AC25 and MYL12B) or anti-correlating (NNT and PARK7) proteoform abundances relative to day 7 sCr.

Next, we performed differential expression analysis to reveal thousands of proteoforms significantly changed within seven days of transplantation (LD: 2206 down-, 660 up-; DCD: 3367 down-, 2140 up-; DBD: 3548 down- and 1739 up-regulated; **Fig. 4d**). Interestingly, while phosphorylation patterns remained similar across recipient groups, deceased donor recipients indicated a shift towards decreased acetylation 7 days post-KTx, where DCD showed higher abundances of acetylated proteoforms at day 0 compared to DBD (**Supplementary Figure S5**). Pathway enrichment analysis of LD versus DCD or DBD revealed a high prevalence of metabolism-associated proteoforms in DCD and LD donors, while cell stress and immune defense were prominent in deceased donor recipients (**Supplementary Figure S6a**). Importantly, we observed distinct upregulation of immune response proteoforms in deceased donors, with the most extreme fold-changes observed in patients who developed an abnormal outcome or rejected the transplant (**Supplementary Figure S6b**). When subjecting only significantly regulated proteoforms to pathway analysis, we observed a distinct profile across all donor groups. While LD recipients significantly upregulated proteoforms involved in wound healing or cell adhesion, deceased donor recipients uniformly upregulated proteoforms involved in homeostasis, platelet activation, and cytoplasmic translation, while cellular component biogenesis was enriched among the significantly downregulated proteoforms. Interestingly, all three donor groups significantly downregulated nucleosome organization and chromatin organization. In-depth follow-up work will be required to explore these pathways and their key contributors (**Fig. 4e**).

When correlating proteoform abundances at day 7 post-KTx with patients’ serum creatinine levels, we identified both positively and negatively correlated proteoforms, many of which were histone proteoforms (**Fig. 4f; Supplementary Figure S6c**). We anticipated that proteoforms exhibiting a trend similar to the clinically widespread sCr evaluation could serve as proteoform biomarkers to distinguish between the donor groups. A closer assessment of blood proteoforms with a q value of less than 0.05 in DCD and DBD but not in LD (total of 174 proteoforms) in correlation to the sCr day 7 levels highlighted acetylated H2AC25 (H2A clustered histone 25, core component of the nucleosome) and MYL12B (myosin light chain 12B, previously associated with septic acute kidney injury) (**Fig. 4g**)^28^. Alternatively, anti-correlating proteoforms, such as NNT (nicotinamide nucleotide transhydrogenase, oxidative stress and mitochondrial dysfunction) and PARK7 (Parkinson disease protein 7, oxidative stress and cell death), were higher in LD compared to the two deceased donor recipients. While most fold changes of proteoforms with a q-value <0.05 showed a similar trend in DCD or DBD (**Supplementary Figure S6d**), sCr-correlating proteoforms showed stark differences between the donor groups. Taken together, these results suggest that donor kidney quality affects the recipient’s blood proteoform landscape, as indicated by differential immunological responses.

### Prioritization of proteoforms identified in deceased donors in relation to discarded kidneys

Finally, to identify which proteoforms could indicate preferential donor quality, we integrated the proteoform differences observed by direct proteoform imaging in LD versus discarded kidney samples. Consistent with the results obtained from LD versus deceased donors, pathway analysis highlighted glycolytic and metabolic pathways in LD over discarded kidneys (**Fig. 1d**). As indicated above, the most differentially expressed proteoform in discarded over transplanted kidneys is monoacetylated CRYAB, a regulator of cell stress, also found significantly upregulated in DCD and DBD over LD donors (**Fig. 1c**, **Fig. 2d-e**). We identified 178 unique proteoforms in the LD donors, 76 in the discarded kidneys, and 28 in both the DBD and DCD donor tissues. As expected, we identified more common unique proteoforms between LD and DCD (23, **Supplementary Figure S7**) compared to DBD (9, **Supplementary Figure S7**) (**Fig. 5a**). Only 4 proteoforms were shared between DBD, DCD and the discarded kidney samples, including the previously indicated acetylated CRYAB (proteoform detailed in **Fig. 5b**), acetylated PARK7 (sensor of oxidative stress), acetylated S100A4 (marker for cell death and stimulator of T-cell chemotaxis) and ACTG1 (cell motility) (**Fig. 5c**). We observed that CRYAB, PARK7 and S100A4 were differentially regulated between LD and the discarded or deceased donors. In contrast, ACTG1 was predominantly observed in the DBD kidneys, even compared with DCD kidneys, indicative of elevated cell motility, actin cytoskeletal reorganization, proliferation, and activation of cellular processes^29^. Therefore, these four proteoform markers largely represent the regulated pathways from our PiMS dataset and could trigger the cell stress response observed in the PBMC data.

**Figure 5.**
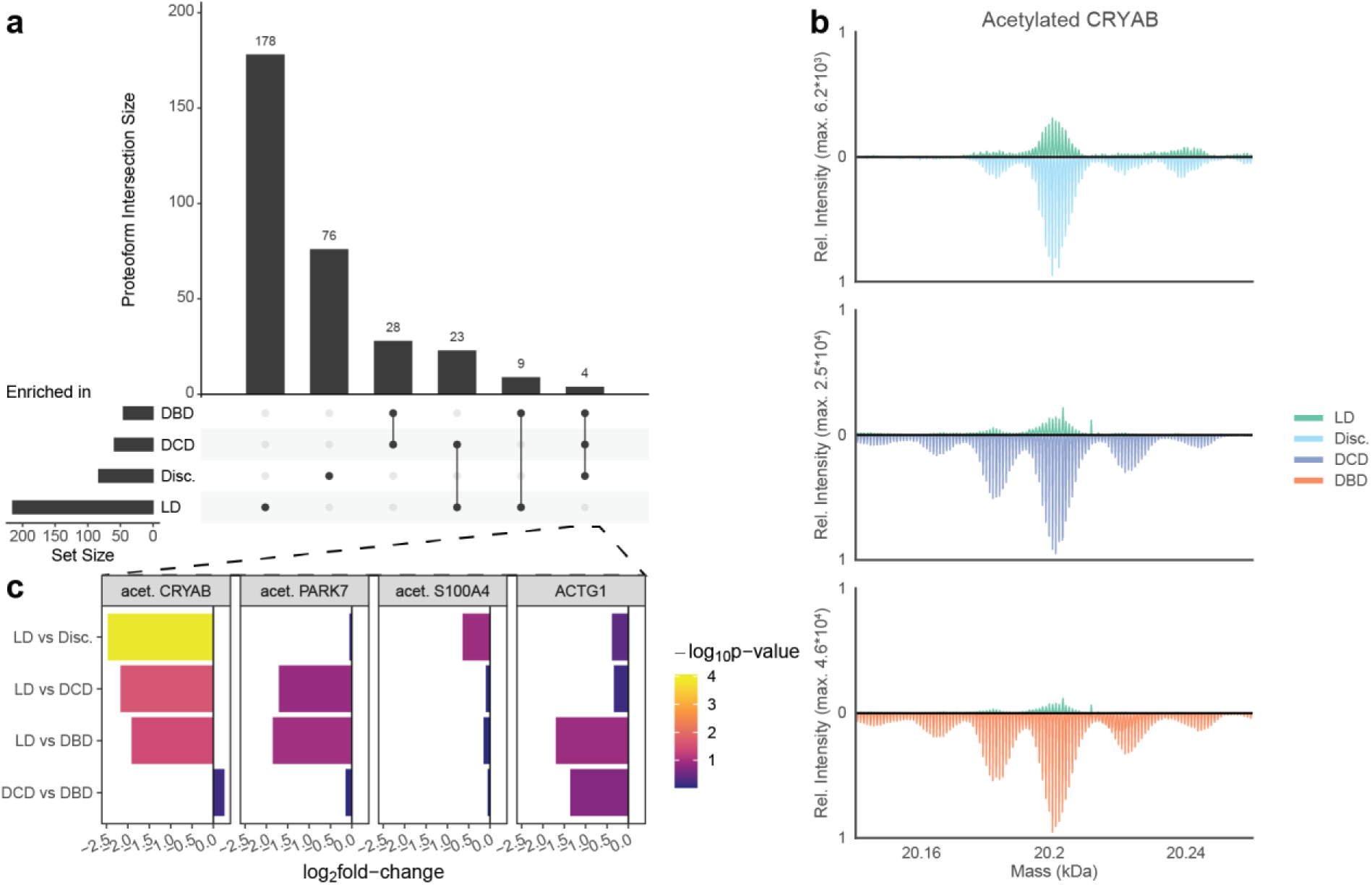
Proteoform overlaps highlight possible markers for tissue quality. **(a)** Upset plot of abundant proteoforms as observed in DBD (versus LD), DCD (versus LD), discarded, Disc. (versus LD), or LD (versus DBD, DCD, and disc.). **(b)** Mass spectra of acetylated CRYAB (20,188 Da) for LD vs. Discarded, DCD, or DBD. Relative intensity with the respective maximum intensity in the indicated comparison and the mass range (kDa) is shown. **(c)** Log_2_ fold-change of 4 shared proteoforms between deceased donors and the discarded kidney across LD vs. Discarded, DCD or DBD, and DCD vs. DBD. Color indicates -log_10_ adjusted p-value.

## Discussion

Reliable molecular markers are crucial to improve quality assessment and the utilization of donor kidneys. Methods to accurately and objectively identify molecular markers associated with metabolic stress, inflammation, and ischemia–reperfusion injury, especially in DBD and DCD, can enhance the evaluation of donor kidney quality, improve utilization, and reduce the ∼30% discard rate of these kidneys. To address this, we used a novel TDP application to characterize spatially-resolved proteoform landscapes and to define an immunometabolic signature in tissue donor biopsies and recipient PBMCs at the systemic level. The common trend between both datasets, dominated by IL-12/JAK-STAT signaling and mitochondrial metabolic remodeling, is consistent with immune activation and cellular stress responses triggered by transplantation, rather than tissue-specific pathology. Specifically, we report the first paired donor tissue and recipient PBMC proteoform profiles for LD, DBD, and DCD kidney transplants. We observed higher levels of metabolic stress markers in deceased-donor kidney tissues and of immune system markers in recipient PBMCs. Integrated proteoform assessment via TDP identifies prospective POC biomarkers that could be used to stratify donor organ quality and improve their utilization, warranting future clinical evaluation in a larger cohort.

It is well established that LD kidneys are considered the gold standard for donor kidney quality, with excellent post-KTx outcomes due to minimal ischemic and metabolic stress and comparatively lower cold ischemic times. We corroborated this using PiMS on tissue biopsies from LD kidneys, where ∼2- to 4-fold higher levels of metabolically active proteoforms were detected. Interestingly, although higher levels of metabolically active proteoforms were also detected in tissue biopsies from DCD kidneys, this could be attributed to metabolic stress or damage that DCD kidneys undergo due to the warm ischemia intrinsic to cardiac death donation (median of 37 vs. 0 minutes in LD donors)^30,31^. As expected, proteoforms associated with cell stress and death, such as acetylated CRYAB and S100A4, were detected in both DBD and DCD kidney biopsies. Deceased donation is known to cause a pro-inflammatory milieu, especially at the endothelial level, resulting in cell stress and death and thus associated with worse post-KTx graft function compared to LD counterparts^32–34^. Interestingly, our TDP results align with existing knowledge of cellular processes in donor kidneys from different donor groups and also provide direct evidence of specific proteoforms involved in these processes, which may serve as candidate biomarkers for tissue quality. It is well established that LD kidneys are considered the gold standard for donor kidney quality, with excellent post-KTx outcomes, due to minimal ischemic and metabolic stress and often low cold ischemic times. We corroborated this using PiMS on tissue biopsies from LD kidneys, where ∼2- to 4-fold higher levels of metabolically active proteoforms were detected. Interestingly, higher levels of metabolically active proteoforms were also detected in tissue biopsies from DCD kidneys, but this could be attributed to metabolic stress or damage that DCD kidneys undergo due to warm ischemia intrinsic to cardiac death donation (37 vs. 0 minutes in LD donors)^30,31^. As expected, proteoforms associated with cell stress and death, such as acetylated CRYAB and S100A4, were detected in both DBD and DCD kidney biopsies. Deceased donation is known to induce a pro-inflammatory milieu, especially at the endothelial level, resulting in cell stress and death, and is thus associated with worse post-KTx graft function compared to LD counterparts^32–34^. Interestingly, our TDP results align with existing knowledge of cellular processes in donor kidneys from different donor groups and also provide direct evidence of specific proteoforms involved in these processes, which may serve as candidate biomarkers for tissue quality.

In addition to detecting local changes in allografts prior to KTx, our comprehensive proteoform analysis of PBMC samples from KTx recipients implanted with LD, DBD, and DCD kidneys reveals that donor kidney quality profoundly influences the recipient’s molecular immune response during the critical first week post-transplantation. While baseline proteoform profiles prior to transplantation show no donor-group-specific clusters, the patient-specific shift at day 7 demonstrates distinct immunological trajectories for LD and deceased donor recipients. The predominance of metabolism-associated proteoforms in LD recipients contrasts with the cell stress and immune defense signatures in deceased donors, suggesting fundamentally different adaptive responses to graft quality. To our knowledge, this is the first study to show a longitudinal difference comparing the graft and systemic immunometabolic signatures across different organ donor types using proteomics. Importantly, the identification of proteoforms correlating with sCr (i.e., acetylated H2AC25 and MYL12B) in deceased donor recipients provides potential molecular biomarkers that may complement traditional clinical assessments. The uniform downregulation of chromatin organization pathways across all donor types, coupled with donor-specific upregulation of homeostasis and platelet activation in deceased donor recipients, highlights the complex interplay between graft quality and systemic immune adaptation. These molecular insights may enable early detection of graft dysfunction, guide immunosuppressive strategies, and improve long-term transplant outcomes.

While analysis of intact proteoform analysis has successfully evaluated donor kidney quality and its impact on KTx^35–37^, we demonstrated here that this more precise type of “top-down” proteomics has several advantages: a) unlike so-called bottom-up proteomics used in previous studies, where intact proteins are digested into peptides, proteoform analysis enables comprehensive and discrete identification of candidate markers in their biologically active forms; b) proteoform profiling of signaling imprinted in combinatorial PTMs to describe the pathophysiological state of the donor kidneys; and c) although not explored in this study, PiMS generates spatially resolved proteoform maps, providing an additional layer of molecular information. Thus, proteoform detection via TDP is now a robust methodology that can provide mechanistic information and help identify molecular markers in small cohorts to assess donor kidney quality and identify early signs of graft rejection. As analysis of intact proteoform analysis has successfully evaluated donor kidney quality and its impact on KTx^35–37^, we demonstrated here that this more precise type of “top-down” proteomics has several advantages: a) unlike so-called bottom-up proteomics used in previous studies, where intact proteins are digested to peptides, proteoform analysis enables comprehensive and discrete identification of candidate markers in their biologically active form; b) proteoform profiling of signaling imprinted in combinatorial PTMs to describe the pathophysiological state of the donor kidneys; and c) although not explored in this study, PiMS generates spatially resolved proteoform maps, providing an additional layer of molecular information. Thus, proteoform detection via TDP is now a robust methodology that can provide mechanistic information and help identify molecular markers in small cohorts to assess donor kidney quality and identify early signs of graft rejection.

The health of donor kidneys is adversely affected during organ procurement, preservation, and implantation, impacting allograft function and survival. Current clinical methods for evaluating donor kidney quality have been in practice for several decades; however, they are subjective and do not fully capture the adverse, multifactorial cellular processes. Detection of intact proteoforms would not only reliably reflect the pathophysiological state of a donor kidney but could also be incorporated into or supplemented by existing evaluation methods to improve donor kidney assessment and utilization. For example, specific proteoforms were detected only in DBD or DCD kidney biopsies. Importantly, we observed coregulation of proteoforms in LD and DBD, or LD and DCD, indicating that, in addition to the clinical data, these proteoforms could serve as objective POC biomarkers of organ viability and predictors of organ utilization and function.

In conclusion, we demonstrate the first comprehensive proteoform landscape of LD, DBD, and DCD kidney biopsies to evaluate donor kidney quality. Additionally, we demonstrate changes in the blood proteoforms in the recipients of LD, DBD, and DCD kidneys post-transplant, highlighting prospective biomarkers and candidate pathways to predict transplantation outcome. Future studies to validate proteoform changes in larger cohorts and longitudinal graft (local) vs. blood (systemic) proteoform correlative analyses will provide an in-depth understanding and characterization of molecular changes, enabling proteoform detection as a clinically actionable biomarker to assess donor kidney quality, advance donor kidney utilization, and predict KTx outcomes.

## Data Sharing Statement

The mass spectrometry datasets generated for this study will be deposited to MASSIVE (https://massive.ucsd.edu/ProteoSAFe/) and will be made available upon publication.

## Acknowledgements

This work was supported by National Institutes of Health grant AI142079-01A1 to S.N.N. Additional funding was received from Northwestern Medicine Dr. Michael M. Abecassis Transplant Innovation Endowment Grant FY2024 to N.L.K. and E.F. We gratefully acknowledge all members of the Kelleher and Nadig laboratories for their support. We acknowledge the use of Biorender to generate the illustrations in Figure 1a.

## Author contributions

S.N.N., D.J., N.L.K., A.S., M.C., and E.F. conceptualized and supervised the project. Cohort and sample acquisition were led by S.N.N., V.R., A.D., D.P., M.A.C., M.E.T., Y.M.L, and D.J. Data were acquired by P.S., N.H., A.S., and C-F.H. Data analysis was performed by I.P., M.H., A.S., M.A.C., and C.C. P.S., A.S., D.J., and C.C. participated in the visualization of the figures. The original draft was written by D.J. and C.C. All authors participated in writing, reviewing, and editing.

## Abbreviations

| Abbreviation | Explanation |
| --- | --- |
| BUN | Blood urea nitrogen |
| CID | Collision-induced dissociation |
| CV | Coefficient of variation |
| DD | Deceased donor |
| DCD | Deceased after cardiac death |
| DBD | Deceased after brain death |
| eGFR | Estimated glomerular filtration rate |
| FDR | False discovery rate |
| GO | Gene ontology |
| I<sup>2</sup>MS | Individual ion mass spectrometry |
| IMT | Intact Mass Tag |
| IS | Immunosuppression |
| IQR | Interquartile range |
| KTx | Kidney transplant |
| KDPO | Kidney Donor Profile Index |
| LC | Liquid chromatography |
| LD | Living Donor |
| MS | Mass spectrometry |
| nano-DESI | Nanospray desorption electrospray ionization |
| PBMCs | Peripheral blood mononuclear cells |
| PEPPI | Passively Eluting Proteins from Polyacrylamide gels as Intact species |
| PiMS | Proteoform imaging mass spectrometry |
| PTM | Post-translational modification |
| sCr | Serum creatinine |
| STORI | Selective Temporal Overview of Resonant Ions |
| SGF | Slow Graft Function |
| TDP | Top-down proteomics |
| TIC | Total ion count |

## References

1. Data & Calculators | HRSA. https://www.hrsa.gov/optn/data-calculators.

2. Lentine, K. L. et al. OPTN/SRTR 2023 Annual Data Report: Kidney. American Journal of Transplantation 25, S22–S137 (2025).

3. Naik, R. H. & Shawar, S. H. Acute Renal Transplantation Rejection. in StatPearls (StatPearls Publishing, Treasure Island (FL), 2025).

4. Kaisar, M. et al. Subclinical Changes in Deceased Donor Kidney Proteomes Are Associated With 12-month Allograft Function Posttransplantation—A Preliminary Study. Transplantation 103, 323 (2019).

5. von Moos, S., Akalin, E., Mas, V. & Mueller, T. F. Assessment of Organ Quality in Kidney Transplantation by Molecular Analysis and Why It May Not Have Been Achieved, Yet. Front. Immunol. 11, (2020).

6. Drown, B. S. et al. Mapping the Proteoform Landscape of Five Human Tissues. J. Proteome Res. 21, 1299–1310 (2022).

7. Aebersold, R. et al. How many human proteoforms are there? Nat Chem Biol 14, 206–214 (2018).

8. Habeck, T. et al. Top-down mass spectrometry of native proteoforms and their complexes: a community study. Nat Methods 21, 2388–2396 (2024).

9. Melani, R. D. et al. The Blood Proteoform Atlas: A reference map of proteoforms in human hematopoietic cells. Science 375, 411–418 (2022).

10. Su, P. et al. Highly multiplexed, label-free proteoform imaging of tissues by individual ion mass spectrometry. Science Advances 8, eabp9929 (2022).

11. Kafader, J. O. et al. Multiplexed mass spectrometry of individual ions improves measurement of proteoforms and their complexes. Nat Methods 17, 391–394 (2020).

12. Forte, E. et al. Top-Down Proteomics Identifies Plasma Proteoform Signatures of Liver Cirrhosis Progression. Mol Cell Proteomics 23, 100876 (2024).

13. Huang, C.-F. et al. Deep Profiling of Plasma Proteoforms with Engineered Nanoparticles for Top-Down Proteomics. J. Proteome Res. 23, 4694–4703 (2024).

14. Huang, C.-F. et al. Targeted Quantification of Proteoforms in Complex Samples by Proteoform Reaction Monitoring. Anal. Chem. 96, 3578–3586 (2024).

15. Zhou, Y. et al. Metascape provides a biologist-oriented resource for the analysis of systems-level datasets. Nat Commun 10, 1523 (2019).

16. Laskin, J., Heath, B. S., Roach, P. J., Cazares, L. & Semmes, O. J. Tissue Imaging Using Nanospray Desorption Electrospray Ionization Mass Spectrometry. Anal. Chem. 84, 141–148 (2012).

17. Denisov, E., Damoc, E. & Makarov, A. Exploring frontiers of orbitrap performance for long transients. International Journal of Mass Spectrometry 466, 116607 (2021).

18. Kafader, J. O. et al. STORI Plots Enable Accurate Tracking of Individual Ion Signals. J. Am. Soc. Mass Spectrom. 30, 2200–2203 (2019).

19. Horn, D. M., Zubarev, R. A. & McLafferty, F. W. Automated reduction and interpretation of High Resolution Electrospray Mass Spectra of Large Molecules. J. Am. Soc. Mass Spectrom. 11, 320–332 (2000).

20. Posit team. RStudio: Integrated Development Environment for R. (Posit Software, PBC, Boston, MA, 2025).

21. Gu, Z. Complex heatmap visualization. iMeta 1, e43 (2022).

22. LeDuc, R. D. et al. Accurate Estimation of Context-Dependent False Discovery Rates in Top-Down Proteomics*[S]. Molecular & Cellular Proteomics 18, i–805 (2019).

23. Ntai, I., Toby, T. K., LeDuc, R. D. & Kelleher, N. L. A Method for Label-Free, Differential Top-Down Proteomics. Methods Mol Biol 1410, 121–133 (2016).

24. Bates, D., Mächler, M., Bolker, B. & Walker, S. Fitting Linear Mixed-Effects Models Using lme4. Journal of Statistical Software 67, 1–48 (2015).

25. Benjamini, Y. & Hochberg, Y. Controlling the False Discovery Rate: A Practical and Powerful Approach to Multiple Testing. Royal Statistical Society. Journal. Series B: Methodological 57, 289–300 (1995).

26. Hulsen, T., de Vlieg, J. & Alkema, W. BioVenn - a web application for the comparison and visualization of biological lists using area-proportional Venn diagrams. BMC Genomics 9, 488 (2008).

27. Humar, A. et al. Effect of initial slow graft function on renal allograft rejection and survival. Clin Transplant 11, 623–627 (1997).

28. Wu, F. et al. Identification of phosphorylated MYL12B as a potential plasma biomarker for septic acute kidney injury using a quantitative proteomic approach. Int J Clin Exp Pathol 8, 14409–14416 (2015).

29. Belyantseva, I. A. et al. γ-Actin is required for cytoskeletal maintenance but not development. Proceedings of the National Academy of Sciences 106, 9703–9708 (2009).

30. Robinson, C. et al. Barriers and Opportunities to Increase Utilization of Donor Kidneys After Death Determined by Circulatory Criteria Among Children and Adults: A Narrative Review. Can J Kidney Health Dis 12, 20543581251382333 (2025).

31. Arnold, M. et al. Metabolic Considerations in Direct Procurement and Perfusion Protocols with DCD Heart Transplantation. International Journal of Molecular Sciences 25, (2024).

32. Taylor, M. E. et al. Mitochondrial responses to brain death in solid organ transplant. Front. Transplant. 2, (2023).

33. Vaughan, R. H. et al. Cytoskeletal protein degradation in brain death donor kidneys associates with adverse posttransplant outcomes. American Journal of Transplantation 22, 1073–1087 (2022).

34. Goncalves Bullock, A. M., Tridente, A. & Dempsey, N. C. Inflammation in deceased kidney donors in the pre-organ retrieval period and the association with transplant outcomes: A systematic review. Transplant Rev (Orlando) 40, 100980 (2025).

35. Huang, C.-F. et al. Mass spectrometry-based proteomics for advancing solid organ transplantation research. Front. Transplant. 2, (2023).

36. Zaza, G. et al. Proteomics reveals specific biological changes induced by the normothermic machine perfusion of donor kidneys with a significant up-regulation of Latexin. Sci Rep 13, 5920 (2023).

37. Hofstraat, R. et al. Highly Repeatable Tissue Proteomics for Kidney Transplant Pathology: Technical and Biological Validation of Protein Analysis using LC-MS/MS. 2024.06.14.599091 Preprint at 10.1101/2024.06.14.599091 (2024).

