## SupplementalFigures for "Top-Down Proteomics in the Assessment of Kidney Donor Quality: a novel approach to increased organ utilization"

Drs. Satish N. Nadig and Neil L. Kelleher

**Running Title**

Top-Down Proteomics for Assessing Kidney Donor Quality

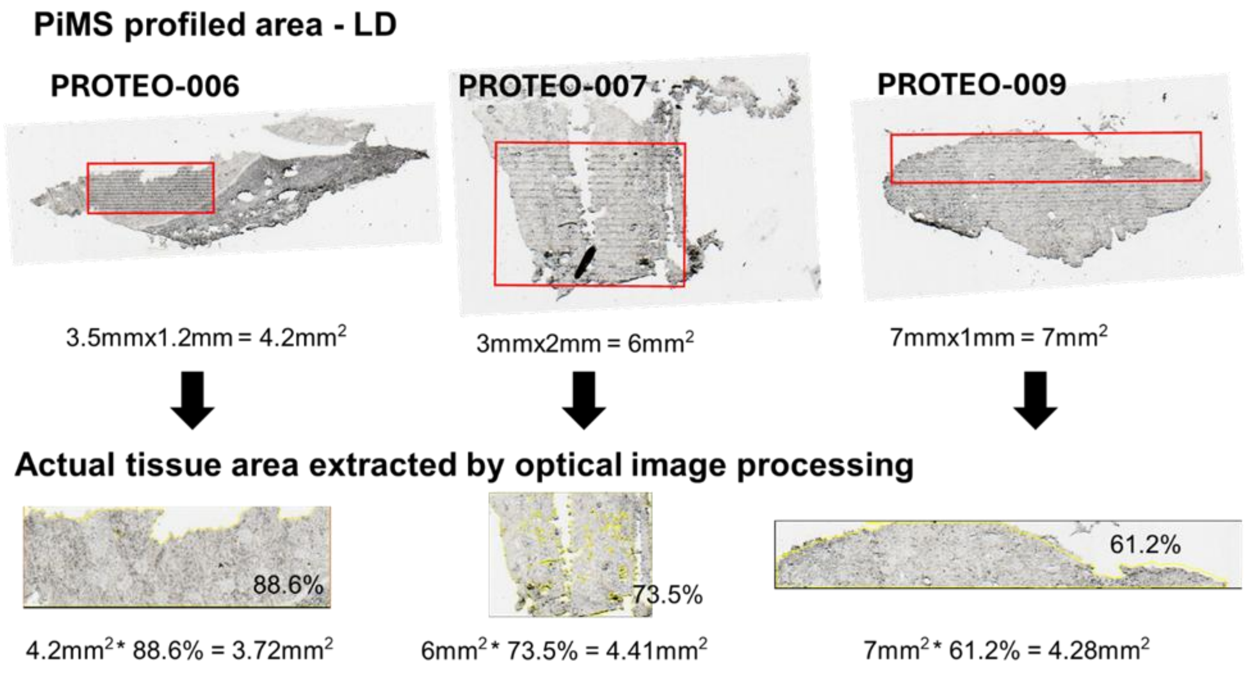


**Supplementary Figure S1.** Optical images of the exact areas selected and the extracted tissue areas for each biopsy in the LD cohort.

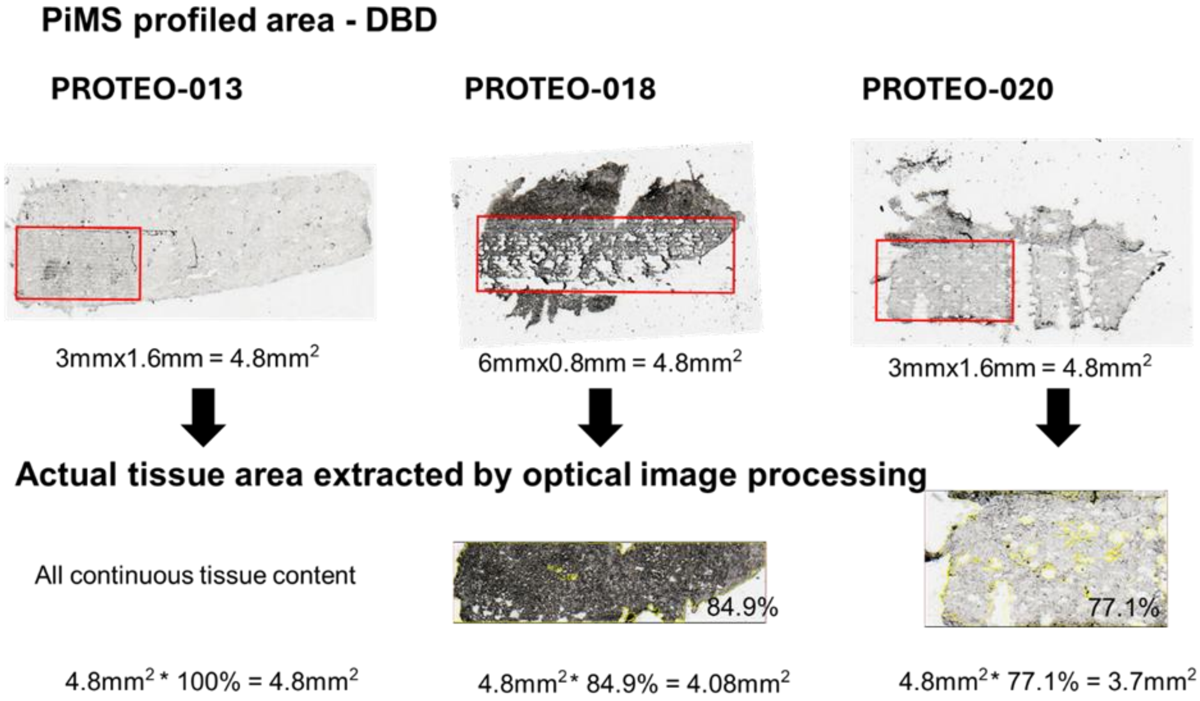


**Supplementary Figure S2.** Optical images of the exact areas selected and the extracted tissue areas for each biopsy in the DBD donor cohort.

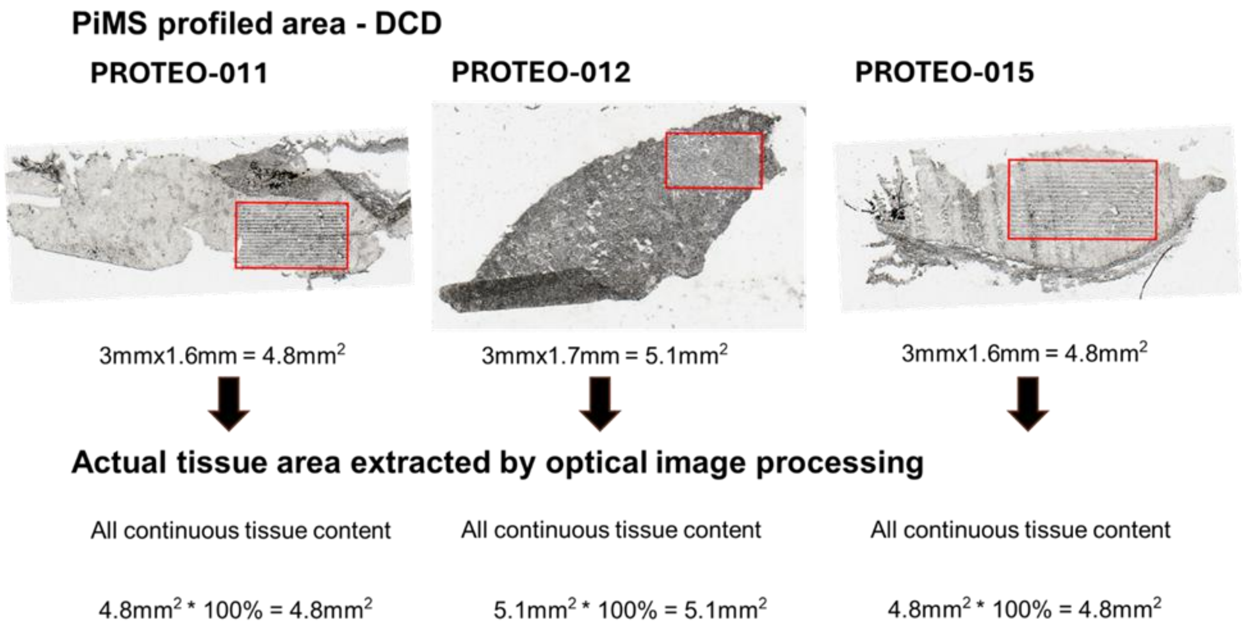


**Supplementary Figure S3.** Optical images of the exact areas selected and the extracted tissue areas for each biopsy in the DCD donor cohort.

| | Overall<br>N = 15 | DBD<br>N = 5 | DCD<br>N = 5 | Living<br>N = 5 | *p-value* |
|---|---|---|---|---|---|
| ***Transplant Information*** | | | **PBMC** | | |
| Kidney Donor Profile Index, % | 58 (40, 63) | 34 (32, 57) | 58 (58, 61) | NA | *0.51* |
| Cold Ischemic Time, minutes | 1,031 (236, 1,319) | 1,075 (1,015, 1,255) | 1,319 (1,175, 1,401) | 234 (219, 235) | *0.06* |
| Warm Ischemic Time, minutes | 36 (28, 43) | 43 (36, 44) | 37 (37, 42) | 28 (26, 29) | *0.14* |
| Perfusion System Use | 5 (56%) | 2 (67%) | 3 (100%) | 0 | *0.14* |
| Resistance | 0.17 (0.17, 0.21) | 0.12 (0.10, 0.15) | 0.21 (0.19, 0.27) | 0 | *0.14* |
| Flow rate, mL/min | 86 (85, 140) | 117 (101, 133) | 86 (86, 113) | 0 | *0.78* |
| Recipient Induction Therapy | | | | | *>0.9* |
| Campath, Steroids | 7 (78%) | 2 (67%) | 2 (67%) | 3 (100%) | |
| Simulect, Steroids | 2 (22%) | 1 (33%) | 1 (33%) | 0 | |
| ***Post-Transplant Outcomes*** | | | | | |
| Slow Graft Function | 4 (44%) | 1 (33%) | 3 (100%) | 0 | *0.14* |
| Dialysis Post-Transplant | 2 (22%) | 1 (33%) | 1 (33%) | 0 | *>0.9* |
| Biopsy Completed | 6 (67%) | 2 (67%) | 2 (67%) | 2 (67%) | *>0.9* |
| Abnormal Result | 3 (50%) | 2 (100%) | 1 (50%) | 0 | *0.6* |
| Rejection | 1 (17%) | 1 (50%) | 0 | 0 | *>0.9* |

**Supplementary Figure S4.** Post-KTx outcomes for all patients included in the PiMS experiments.

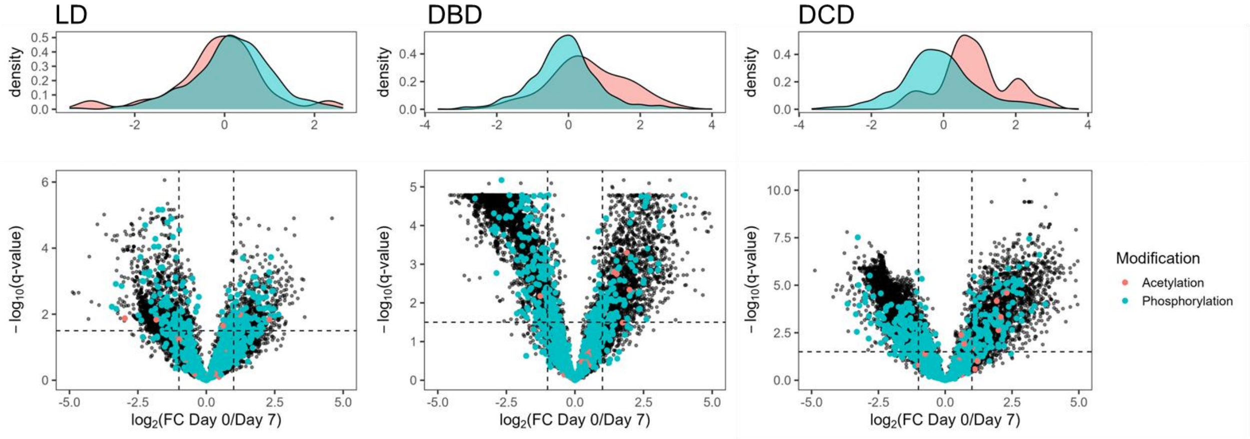


**Supplementary Figure S5.** $Log_2$ fold-change of LD, DBD, and DCD for pre- and day 7 post- KTx and $-\log_{10}$ q-value indicating acetylation and phosphorylation patterns.

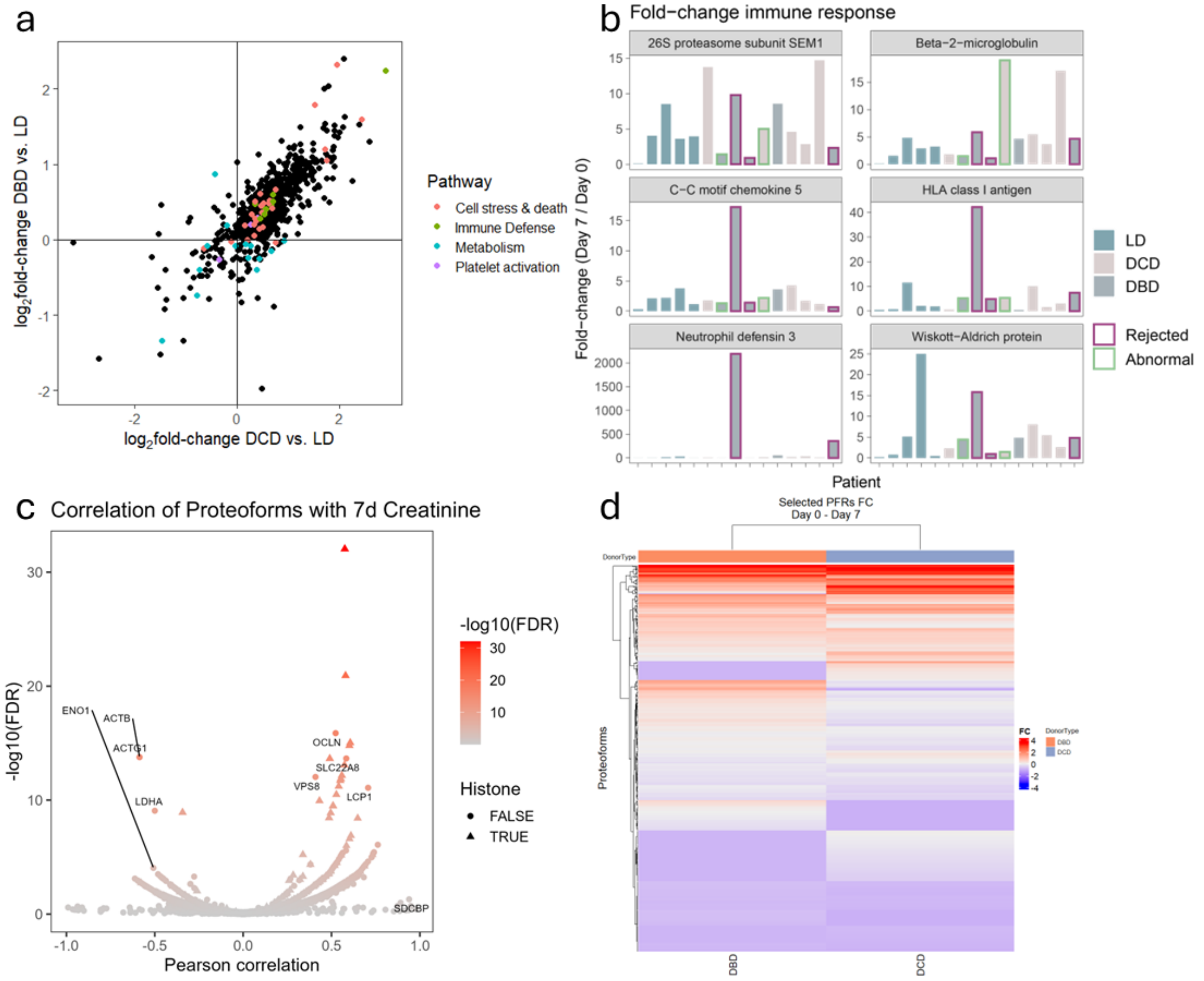


**Supplementary Figure S6. (a)** 2D enrichment of PBMC proteoform fold-changes (day 0 to day 7) LD relative to DCD (x-axis) or DBD (y-axis). Selected pathways are indicated for cell stress and death, platelet activation, immune defense, and metabolism (summarized pathways are detailed in supplementary methods). **(b)** Fold change of selected immune response-associated proteoforms for LD, DCD and DBD recipients, border color indicates patient outcome. **(c)** Pearson correlation of proteoform abundances with sCr at day 7 across LD, DCD, and DBD recipients. Colors according to -log$_{10}$ adjusted p-value and triangles indicate histones. **(d)** Fold-changes of proteoforms with q-value <0.05 in DBD, DCD, and not LD.

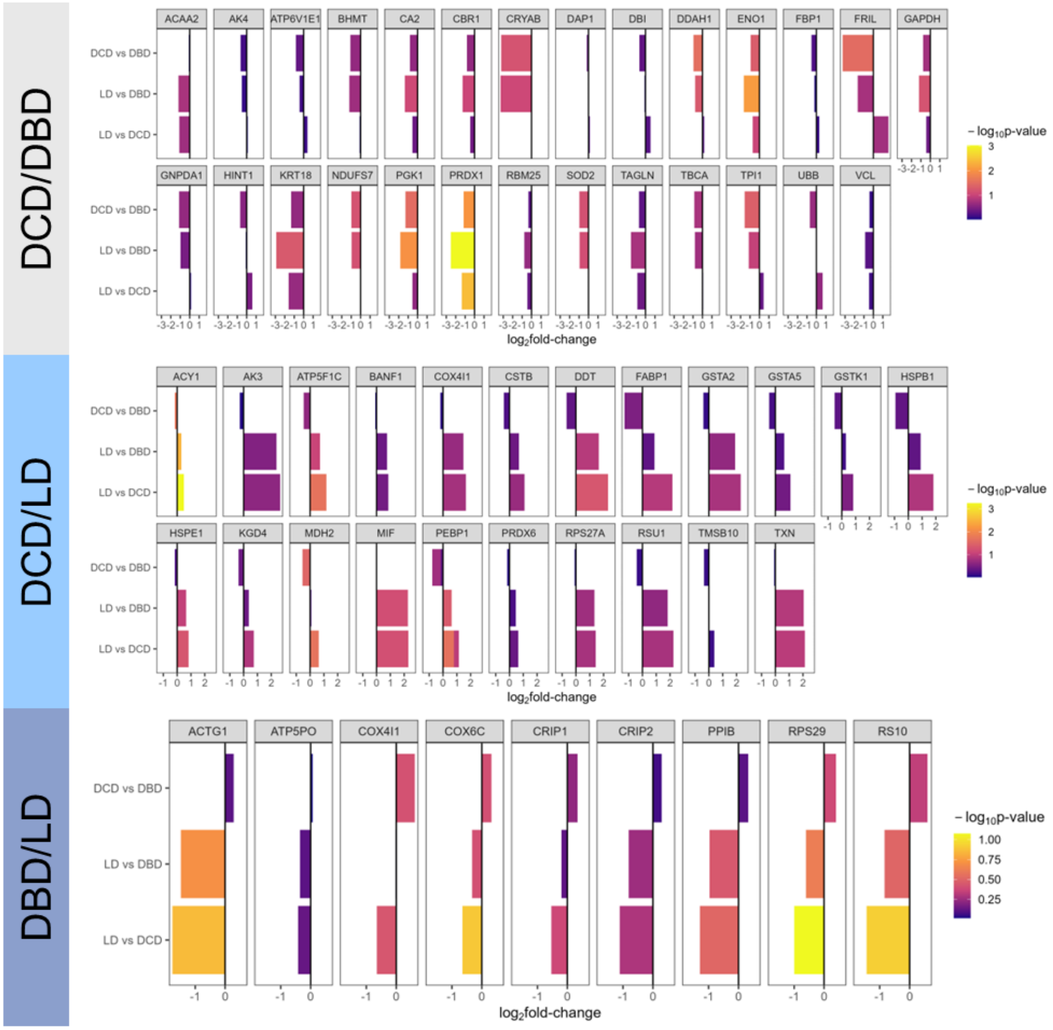


**Supplementary Figure S7.** Log$_2$ fold-changes across DCD versus DBD, LD versus DBD or DCD for proteoforms exclusively identified in DCD and DBD, DCD and LD, or DBD and LD. Colors indicate -log$_{10}$ p-value.
